# Leisure-time physical activity, sedentary behavior, and biological aging: evidence from genetic correlation and Mendelian randomization analyses

**DOI:** 10.1101/2024.07.25.24310997

**Authors:** Xunying Zhao, Xueyao Wu, Lin He, Jinyu Xiao, Rong Xiang, Linna Sha, Mingshuang Tang, Yu Hao, Yang Qu, Changfeng Xiao, Chenjiarui Qin, Jiaojiao Hou, Qin Deng, Jiangbo Zhu, Sirui Zheng, Jinyu Zhou, Ting Yu, Bin Yang, Xin Song, Tao Han, Jiaqiang Liao, Tao Zhang, Mengyu Fan, Jiayuan Li, Xia Jiang

**Affiliations:** Department of Epidemiology and Health Statistics and West China-PUMC C. C. Chen Institute of Health, West China School of Public Health and West China Fourth Hospital, Sichuan University, Chengdu, Sichuan, China; Division of Cancer Epidemiology and Genetics, National Cancer Institute, Rockville, Maryland, USA; Department of Nutrition and Food Hygiene, West China School of Public Health and West China Fourth Hospital, Sichuan University, Chengdu, China; Department of Clinical Neuroscience, Center for Molecular Medicine, Karolinska Institutet, Solna, Stockholm, Sweden

**Keywords:** physical activity, sedentary behavior, biological aging, epigenetic clock, Mendelian randomization, causal inference, genetic correlation

## Abstract

Physical inactivity and sedentary behavior are associated with higher risks of age-related morbidity and mortality. However, whether they causally contribute to accelerating biological aging has not been fully elucidated. Utilizing the largest available genome-wide association study (GWAS) summary data, we implemented a comprehensive analytical framework to investigate the causal relationships between moderate-to-vigorous leisure-time physical activity (MVPA), leisure screen time (LST), and four epigenetic age acceleration (EAA) measures: HannumAgeAccel, intrinsic HorvathAgeAccel, PhenoAgeAccel, and GrimAgeAccel. Shared genetic backgrounds across these traits were quantified through genetic correlation analysis. Overall and independent causal effects were assessed through univariable and multivariable Mendelian randomization (MR). A recently developed tissue-partitioned MR approach was further adopted to explore potential tissue-specific pathway that contributes to the observed causal relationships. Among the four EAA measures investigated, consistent results were identified for PhenoAgeAccel and GrimAgeAccel. These two measures were negatively genetically correlated with MVPA (*r*_*g*_=−0.18∼−0.29) and positively genetically correlated with LST (*r_*g*_*=0.22∼0.37). Univariable MR yielded a robust effect of genetically predicted LST on GrimAgeAccel (*β*_IVW_ =0.69, *P*=1.10×10^−7^), while MVPA (*β*_IVW_=−1.02, *P*=1.50×10^−2^) and LST (*β*_IVW_=0.37, *P*=1.90×10^−2^) showed marginal causal effects on PhenoAgeAccel. Multivariable MR suggested an independent causal role of LST in GrimAgeAccel after accounting for effects of MVPA and other important confounders. Tissue-partitioned MR suggested skeletal muscle tissue associated variants be predominantly responsible for driving the effect of LST on GrimAgeAccel. Findings support sedentary lifestyles as a modifiable causal risk factor in accelerating epigenetic aging, emphasizing the need for preventive strategies to reduce sedentary screen time for healthy aging.

## Introduction

Aging is a dynamic and intricate process characterized by the gradual accumulation of perturbations, spanning from subtle molecular changes to observable alterations in physiology and functionality^1^. Due to the heterogeneity observed in the aging processes among individuals of similar chronological age^2^, biological age taking into account the internal physiological states and inter-individual variations represents a more precise indicator of aging^3^. Epigenetic clocks, which estimate chronological age or related phenotypes (e.g., mortality) based on levels of DNA methylation (DNAm) at specific cytosine-phosphate-guanine (CpG) sites^4^, have emerged as one of the most promising measures of biological aging^5^. Accumulating evidence suggests that exceeding chronological age according to epigenetic clock estimates, known as epigenetic age acceleration (EAA), is linked to multiple adverse aging-related health consequences^6–8^.

Epigenetic clocks are largely modifiable^9^, identifying intervenable factors such as lifestyle choices that can decelerate or reverse EAA thus holds great potential in informing early interventions to promote healthy aging. Among them, physical activity and sedentary behavior are believed to play a role in reducing age-related morbidity and enhancing lifespan^10–12^. While several epidemiological studies have investigated the “physical activity-EAA” link, results across different epigenetic clocks remain inconsistent. For instance, a study involving 284 participants reported a reverse association between daily step counts with extrinsic Hannum age acceleration (HannumAgeAccel, *β* = −0.100, *P* = 0.027), but a positive association between sit-to-stand transitions with intrinsic Horvath age acceleration (HorvathAgeAccel, *β* = 0.006, *P* = 0.049)^13^. However, two subsequent studies, one involving 3,567 participants and the other involving 2,758 participants, found no evidence to support such associations^14^ ^15^. Meanwhile, the putative impact of sedentary behavior on biological aging remains largely unexplored, despite the recognition that its adverse health effects may largely be independent of physical activity levels^16^ ^17^. In addition to the potential influence of insufficiently active lifestyles on aging, the aging process itself, accompanied by declining physical fitness, may also contribute to changes in physical activity and sedentary behavior^18^. Owing to biases derived from confounding and reverse causality, it is therefore difficult for conventional epidemiological studies to elucidate the causal relationships.

One way to evaluate causality while mitigating the limitations of conventional epidemiological studies is through Mendelian randomization (MR), a framework that utilizes genetic variants (single-nucleotide polymorphism, SNP) as instrumental variables (IVs) to make causal inference by fulfilling three key assumptions (the relevance assumption, the independence assumption, and the exclusion restriction assumption)^19^. Multivariable MR extends this approach to assess the independent causal effects of multiple exposures on an outcome^20^. Further, tissue-partitioned MR that builds upon multivariable MR separates the phenotypic subcomponents of an exposure, allowing for the identification of tissue-specific subcomponent that predominantly drives its causal effect on an outcome, providing insights into the underlying biological mechanisms^21^ ^22^. To the best of our knowledge, no MR study of sedentary behavior with EAA has been performed; only one existing MR assessed the putative effect of physical activity on EAA and identified no evidence of a causal relationship using the then available IVs (N = 6)^23^. Given the discovery of additional genetic loci for each trait of interest^24^ ^25^ and recent developments in novel methodological techniques, a comprehensive MR study is urgently needed to validate and expand previous findings.

Here, leveraging the hitherto largest genome-wide association study (GWAS) summary statistics and comprehensive genetic correlation and MR analyses, we aimed to (i) understand the average shared genetic basis underlying physical activity, sedentary behavior and EAA measures; (ii) investigate the overall causal effects of genetically predicted physical activity and sedentary behavior on each EAA measure; (iii) examine their independent causal effects after accounting for the interplay between physical activity and sedentary behavior, as well as the confounding effects from other important factors; (iv) explore which tissue-dependent biological pathways may predominantly contribute to the observed causal effects.

## Methods

In our study, we used publicly available summary-level data that had obtained ethical approval in all original studies. We followed the guidelines of the Strengthening the Reporting of Observational Studies in Epidemiology - Mendelian Randomization (STROBE-MR)^26^ (**Supplementary File 1,** https://www.strobe-mr.org/). Flowchart of the overall study design is depicted in **Figure 1**.

**Figure 1.**
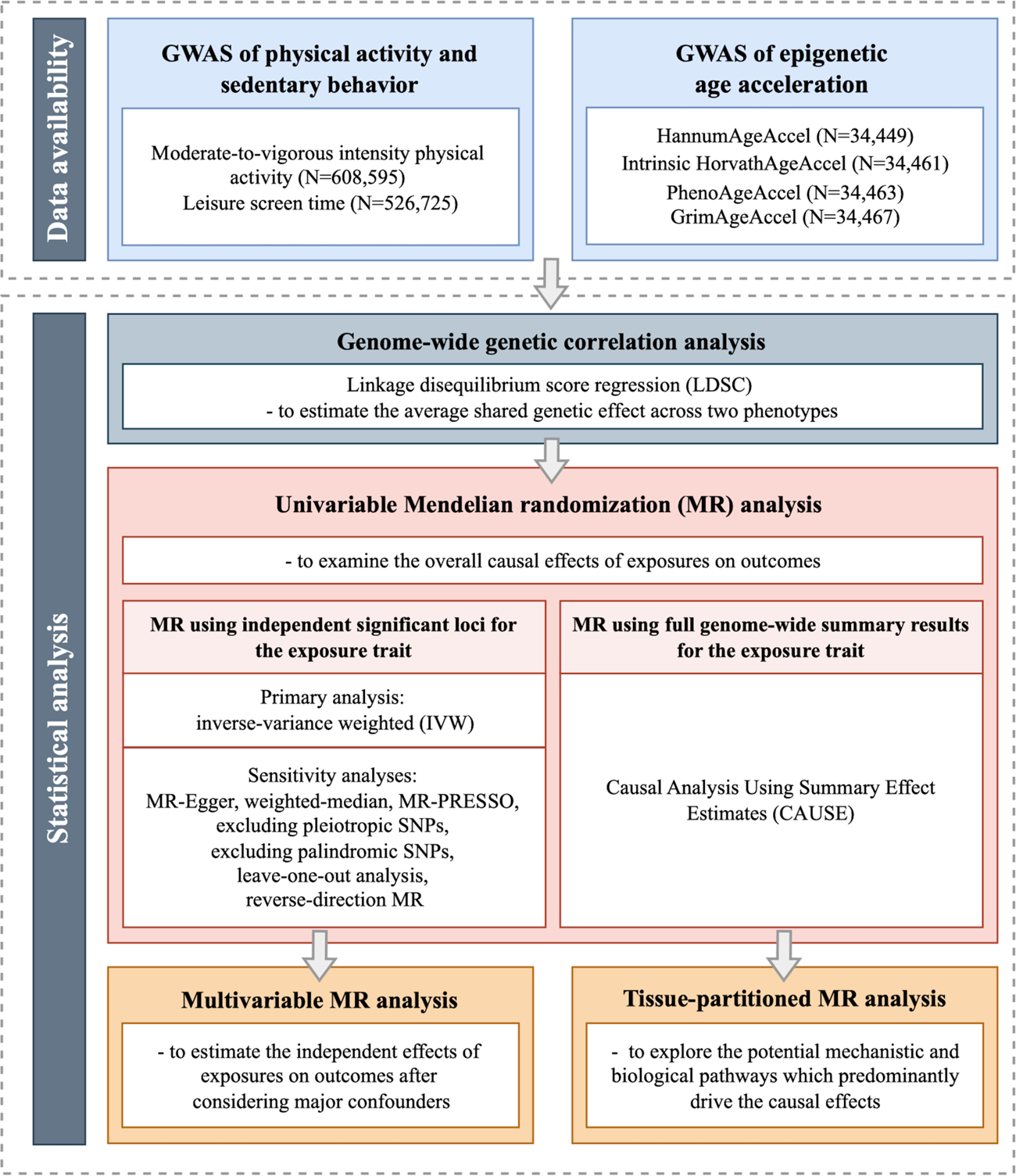
The flowchart of the study. GWAS (genome-wide association study), HannumAgeAccel (Hannum age acceleration), Intrinsic HorvathAgeAccel (intrinsic Horvath age acceleration), PhenoAgeAccel (PhenoAge acceleration), and GrimAgeAccel (GrimAge acceleration).

### GWAS datasets

#### Physical activity and sedentary behavior

We obtained the hitherto largest summary-level data for physical activity and sedentary behavior from a GWAS meta-analysis of 51 studies, comprising 661,399 participants of European ancestry^24^. In brief, this meta-analysis utilized questionnaire-based data, capturing self-reported domain- and intensity-specific physical activity and sedentary traits as phenotypes. Moderate-to-vigorous leisure-time physical activity (MVPA) and leisure screen time (LST) were used as proxies for levels of leisure-time physical activity and sedentary behavior, respectively. MVPA (N = 608,595, SNP-heritability = 8%) was categorized as a dichotomous variable to account for the heterogeneity of the questionnaires used across cohorts, whereas LST (N = 526,725, SNP-heritability = 16%) was defined as a continuous variable.

#### Epigenetic clock acceleration

We obtained the hitherto largest summary-level data for EAA measures from a GWAS meta-analysis of 30 cohorts, comprising 34,710 participants of European ancestry^25^. Four EAA measures were chosen to index epigenetic age acceleration, including HannumAgeAccel^27^ (N = 34,449), intrinsic HorvathAgeAccel^28^ (N = 34,461), PhenoAge acceleration^29^ (PhenoAgeAccel, N = 34,463), and GrimAge acceleration^30^ (GrimAgeAccel, N = 34,467), with SNP-heritability estimated ranging from 10% to 17%.

The Hannum and Horvath clocks, referred to as the first-generation epigenetic clocks, were developed to estimate chronological age based on DNAm data from blood and human tissues/cell types, respectively^27^ ^28^. Intrinsic HorvathAgeAccel, as a derivative of the Horvath clock, was adjusted to exclude the effects of blood cell composition and capture the cell-intrinsic properties of aging^31^. PhenoAge and GrimAge, recognized as the second-generation epigenetic clocks, were developed to predict health and lifespan by incorporating DNAm information associated with mortality and certain clinical biomarkers^29^ ^30^. Details of these EAA measures can be found in the referenced publications^25^ ^27–30^.

#### Confounders

Six additional factors, including educational attainment, smoking, drinking, obesity, sleep duration, and parental longevity, were considered potential confounders in the MVPA/LST-EAA relationship. We obtained GWAS summary data for educational attainment from the Social Science Genetic Association Consortium, involving 766,345 individuals of European ancestry after excluding subjects of 23andMe (due to data restrictions)^32^. For smoking and drinking, we obtained GWAS summary data from GWAS & Sequencing Consortium of Alcohol and Nicotine, involving 632,802 individuals of European ancestry for smoking initiation and 537,349 individuals of European ancestry for alcohol consumption (both excluded subjects of 23andMe)^33^. For obesity, we used GWAS summary data from a GWAS meta-analysis of the UK Biobank and the Genetic Investigation of Anthropometric Traits Consortium, involving 806,834 individuals of European ancestry for body-mass index (BMI)^34^. For sleep duration and parental longevity, we used GWAS summary data of European ancestry from the UK Biobank, including data on sleep duration for 446,118 individuals and on parental age at death for 208,118 individuals, respectively^35^ ^36^.

Details on the characteristics of each GWAS dataset are presented in **Supplementary Table 1.**

### Statistical analysis

#### Genome-wide genetic correlation

We first conducted a genome-wide genetic correlation analysis to quantify the average shared genetic effects underlying MVPA, LST and EAA measures using the linkage-disequilibrium (LD) score regression (LDSC) software^37^. The genetic correlation estimates (*r_*g*_*) range from −1 to +1, where +1 denotes a total positive correlation and −1 denotes a total negative correlation. We used pre-computed LD-scores of ∼1.2 million common SNPs in European ancestry from the Hapmap3 reference panel, aligning with the European origin of our GWAS samples. According to Bonferroni correction, a statistically significant *r_*g*_* was defined as *P* < 6.25×10^−3^ (α = 0.05/8, number of trait-pairs), and a suggestively significant *r_*g*_* was defined as 6.25×10^−3^ ≤ *P* < 0.05^38^.

#### Univariable Mendelian randomization

To investigate the overall causal effects of genetically predicted MVPA or LST on EAA measures, a univariable MR was first performed through exposure-associated SNPs as IVs, then through the genome-wide summary statistics for the exposure traits.

To meet the relevance assumption of MR, we screened IVs through a clumping strategy of *P* < 5×10^-8^ and an LD window of ± 1.0 Mb (*r^g^* < 0.001). For IVs that were not available in the outcome GWAS, we used LDlinkR (R package) to identify proxy SNPs in LD (*r^g^*> 0.8) with the index SNPs from the outcome GWAS data^39^. We calculated the proportion of trait variance explained by each IV (*R^2^*), as well as their *F*-statistics. An *F*-statistic below 10 suggests a weak instrument, which would be excluded from the analysis^40^. Statistical power was calculated with a web-based application (https://sb452.shinyapps.io/power/)^41^. Heterogeneity of the instruments was calculated using Cochran’s Q statistics^42^. Details on the characteristics of IVs are presented in **Supplementary Table 2-3.**

We applied inverse-variance weighted (IVW) as our primary approach. This method pools the estimate from each IV and provides an overall estimate of the causal effect assuming all IVs to be valid (meet the MR assumptions); or are invalid in such a way that the overall pleiotropy is balanced to be zero^19^ ^41^. We performed several sensitivity analyses to assess the robustness of the primary results and to further validate the exclusion restriction and independence assumptions of MR^41^. These include: (i) MR-Egger regression incorporating an intercept term to account for directional pleiotropy^43^; (ii) weighted-median approach exhibiting stronger robustness against invalid IVs^44^; (iii) MR-Pleiotropy Residual Sum and Outlier (MR-PRESSO) dealing with uncorrelated pleiotropy based on outlier removal^45^; (iv) IVW excluding palindromic IVs with strand ambiguity; (v) IVW excluding pleiotropic IVs associated with potential confounders (accessed in GWAS catalog on 02/20/2024, https://www.ebi.ac.uk/gwas/; pleiotropic IVs are listed in **Supplementary Table 2**); (vi) leave-one-out analysis where each IV was excluded at a time and IVW was conducted using the remaining IVs; (vii) IVW-based reverse-direction MR to rule out the possibility of causal effects of EAA measures on MVPA or LST.

We then employed the Causal Analysis Using Summary Effect Estimates (CAUSE) method^46^. Integrating information from genome-wide SNPs, CAUSE has several advantages over the conventional MR approaches: it accounts for both uncorrelated and correlated horizontal pleiotropy, improves statistical power, corrects for potential sample overlap, and reduces the likelihood of false positives. Under a Bayesian framework, CAUSE assumes a proportion (*q* value) of variants that are likely to show correlated horizontal pleiotropy, and provides posterior distribution estimates under two models - the sharing model, which allows only for horizontal pleiotropic effects, and the causal model, which accommodates both horizontal pleiotropy and causality. A one-sided *P*_causal vs. sharing_ is generated to evaluate whether the sharing model is at least as effective as the causal model in fitting the data, with a rejection of the null hypothesis (*P*_causal vs. sharing_ < 0.05) indicating that the data are more likely to be explained by causal effects. To mitigate the impact of strong regional LD structure, we excluded variants within the Major Histocompatibility Complex (MHC) region (chr6: 25M-35M).

Bonferroni correction was applied across all univariable MR approaches, considering a *P* < 6.25×10^−3^ as evidence for statistical significance and a 6.25×10^−3^ ≤ *P* < 0.05 as suggestive significance^38^. To define a robust causal effect, we required that the MR effect estimate demonstrated statistical significance in any univariable MR approach and remained directionally consistent across all approaches, and its null hypothesis for model fitting was rejected (*P*_causal vs. sharing_ < 0.05) in CAUSE.

#### Multivariable Mendelian randomization

To further evaluate the independent causal effect of MVPA or LST on each EAA measure, a multivariable MR was subsequently conducted^20^. Given the potential inter-correlation between MVPA and LST, a multivariable model (referred to as Model 1) was first constructed incorporating both MVPA and LST to estimate their causal effects on EAA independent from each other. Considering potential confounding effects from other factors, a second multivariable model (Model 2) was developed, wherein each speculated confounding factor (i.e., educational attainment, smoking, drinking, BMI, sleep duration, and parental age at death) was included individually together with the exposure. In the third multivariable model (Model 3), confounders in Model 2 were included simultaneously to assess their combined influence. We removed SNPs in LD (*r^g^*≥ 0.001) to avoid overlapping or correlated SNPs after combining different sets of IVs (**Supplementary Table 4-7**). Conditional *F*-statistics were calculated to evaluate the joint instrument strength in multivariable MR settings, with values below 10 indicating weak instruments^47^. According to Bonferroni correction, a *P* < 6.25×10^−3^ was considered as evidence for statistical significance, and a 6.25×10^−3^ ≤ *P* < 0.05 as suggestive significance^38^.

#### Tissue-partitioned Mendelian randomization

Previous post-GWAS analyses have shown that loci associated with MVPA or LST are mainly enriched for gene expressions in brain and skeletal muscle tissues^24^. Both brain and skeletal muscle tissues play a non-negligible role in regulating physical activity and sedentary behavior – while the brain is responsible for generating and regulating behavioral patterns and motivation, the skeletal muscle facilitates or restricts movement^48^. Hence, MVPA/LST influencing genes expressed in the brain may be more likely to achieve their effects by regulating behavior, whereas those expressed in skeletal muscle may impact greater extent pathways related to muscle metabolism. Accordingly, the observed causal effects of MVPA/LST on EAA may involve separate pathways mediated through the brain or skeletal muscle. While other pathways may also likely mediate these effects, they are not the most relevant candidates in this study.

To investigate the primary biological pathways underlying the observed causal relationships, we finally performed an exploratory tissue-partitioned MR. This approach enables us to separate the effects of phenotypic subcomponents of MVPA/LST, here, the “brain-tissue instrumented MVPA/LST” and the “skeletal muscle-tissue instrumented MVPA/LST”, through fractionation of the original IVs according to whether they colocalize with gene expression in the brain or skeletal muscle tissue^21^.

Specifically, at each locus surrounding the original MVPA/LST IVs (within a 200 kb window), we used the Bayesian method “*coloc*” to assess the presence of a single causal variant responsible for both the MVPA/LST GWAS signal and the tissue-specific gene-expression-association signal, a phenomenon referred to as colocalization. Colocalization analyses were conducted twice at each locus: first with expression quantitative trait loci (eQTL) data derived from brain tissue and then separately with eQTL data from skeletal muscle tissue. A posterior probability (PPH4) ≥ 0.8 was considered as strong evidence of colocalization, as recommended by authors of the original method^21^ ^49^. Consequently, the initial set of IVs was divided into two sets, with each set specifically indexing the brain- or the skeletal muscle-tissue instrumented MVPA/LST (tissue-specific IVs). We obtained the brain eQTL dataset from a meta-analysis study of 10 brain regions (N = 1,194, all of European ancestry)^50^, and the skeletal muscle eQTL dataset from the GTEx consortium v.8 (N = 706, ∼80% European ancestry)^51^. Variants residing within the MHC region (chr6: 25M-35M) were excluded from the analysis.

Exposure-outcome pairs that showed robust causal relationships in univariable MR and directionally consistent estimates in multivariable MR were selected for inclusion in the tissue-partitioned analysis. Utilizing the derived tissue-specific IVs, we first performed a univariable MR to evaluate the unadjusted effect of each phenotypic subcomponent of the exposure(s) on the outcome(s). Following this, a multivariable MR was performed to estimate the independent effect of each subcomponent, with IVs weighted using their tissue-specific PPH4 values^21^. Given the largely reduced number of IVs and the resulting decreased statistical power, we used a conventional significance threshold of *P* < 0.05 in the analysis. Further details regarding the analytical procedures can be found in **Supplementary File 2**.

All MR analyses were conducted using R software (v4.1.0) with packages including “TwoSampleMR” (v0.5.4), “MRPRESSO” (v1.0), “MendelianRandomization” (v0.7.0), “CAUSE” (v1.2.0), “MVMR” (v0.4), and “coloc” (v5.1.0).

## Results

### Genome-wide genetic correlation

After Bonferroni correction, we observed statistically significant negative genetic correlations of MVPA with PhenoAgeAccel (*r_g_* = −0.18, *P* = 2.40×10^−3^) and GrimAgeAccel (*r_g_* = −0.29, *P* = 9.69×10^−7^), as well as statistically significant positive genetic correlations of LST with PhenoAgeAccel (*r_g_* = 0.22, *P* = 1.02×10^−5^) and GrimAgeAccel (*r_g_* = 0.37, *P* = 1.14×10^−11^). A suggestively significant genetic correlation was observed for LST with HannumAgeAccel (*r_g_* = 0.08, *P* = 4.84×10^−2^). No significant genetic correlations were found for other trait-pairs (Table 1).

**Table 1.** Genetic correlation between MVPA and LST with each measure of EAA.

| **Triat1** | **Trait2** | **$r_g$** | **95%CI** | **$P$-value** |
| --- | --- | --- | --- | --- |
| MVPA | HannumAgeAccel | −0.073 | (−0.186, 0.040) | 0.20 |
|  | Intrinsic HorvathAgeAccel | −0.017 | (−0.116, 0.083) | 0.74 |
|  | **PhenoAgeAccel** | **−0.179** | **(−0.294, −0.063)** | **$2.40 \times 10^{-3}$** |
|  | **GrimAgeAccel** | **−0.294** | **(−0.412, −0.177)** | **$9.69 \times 10^{-7}$** |
| LST | HannumAgeAccel | 0.084 | (0.001, 0.168) | $4.84 \times 10^{-2}$ |
|  | Intrinsic HorvathAgeAccel | 0.070 | (−0.001, 0.142) | 0.05 |
|  | **PhenoAgeAccel** | **0.222** | **(0.123, 0.320)** | **$1.02 \times 10^{-5}$** |
|  | **GrimAgeAccel** | **0.368** | **(0.262, 0.474)** | **$1.14 \times 10^{-11}$** |

Bold-face: $P$ -value $< 6.25 \times 10^{-3}$ . MVPA (Moderate-to-vigorous intensity physical activity during leisure time), LST (leisure screen time), EAA (epigenetic age acceleration), HannumAgeAccel (Hannum age acceleration), Intrinsic HorvathAgeAccel (intrinsic Horvath age acceleration), PhenoAgeAccel (PhenoAge acceleration), GrimAgeAccel (GrimAge acceleration), $r_g$ (genetic correlation), and 95%CI (95% confidence interval).

### Univariable Mendelian randomization

Motivated by the significant genetic overlap, we proceeded to examine the potential causal effects of MVPA and LST on EAA measures. Altogether 15 independent SNPs were determined as IVs for MVPA, and 130 independent SNPs as IVs for LST. *F*-statistics of these IVs indicated a minimal likelihood of weak instrument bias (all *F*-statistics > 10; **Supplementary Table 2-3**). At an alpha level of 0.05, our univariable MR was estimated to have 80% power to detect causal estimates (*β*) ranging from 0.15 to 0.50 **(Supplementary Table 8)**.

As shown in **Figure 2**, we identified a statistically significant relationship between genetically predicted longer LST and faster GrimAgeAccel (*β*_IVW_ = 0.69, 95% confidence intervals, 95%CIs = 0.43 ∼ 0.94, *P* = 1.10×10^−7^; **Supplementary Table 9**). Despite a modest heterogeneity indicated across individual SNP estimates (*P*_Cochran’s Q_ = 0.02), all sensitivity analyses generated statistically significant and directionally consistent results **(Supplementary Table 9-11)**. We also found that genetically predicted lower levels of MVPA (*β*_IVW_ = −1.02, 95%CIs = −1.84 ∼ −0.20, *P* = 1.50×10^−2^) and genetically predicted longer LST (*β*_IVW_ = 0.37, 95%CIs = 0.06 ∼ 0.67, *P* = 1.90×10^−2^) were associated with faster PhenoAgeAccel under suggestive significance. No evidence of a significant causal effect of MVPA or LST on HannumAgeAccel or intrinsic HorvathAgeAccel was found.

**Figure 2.**
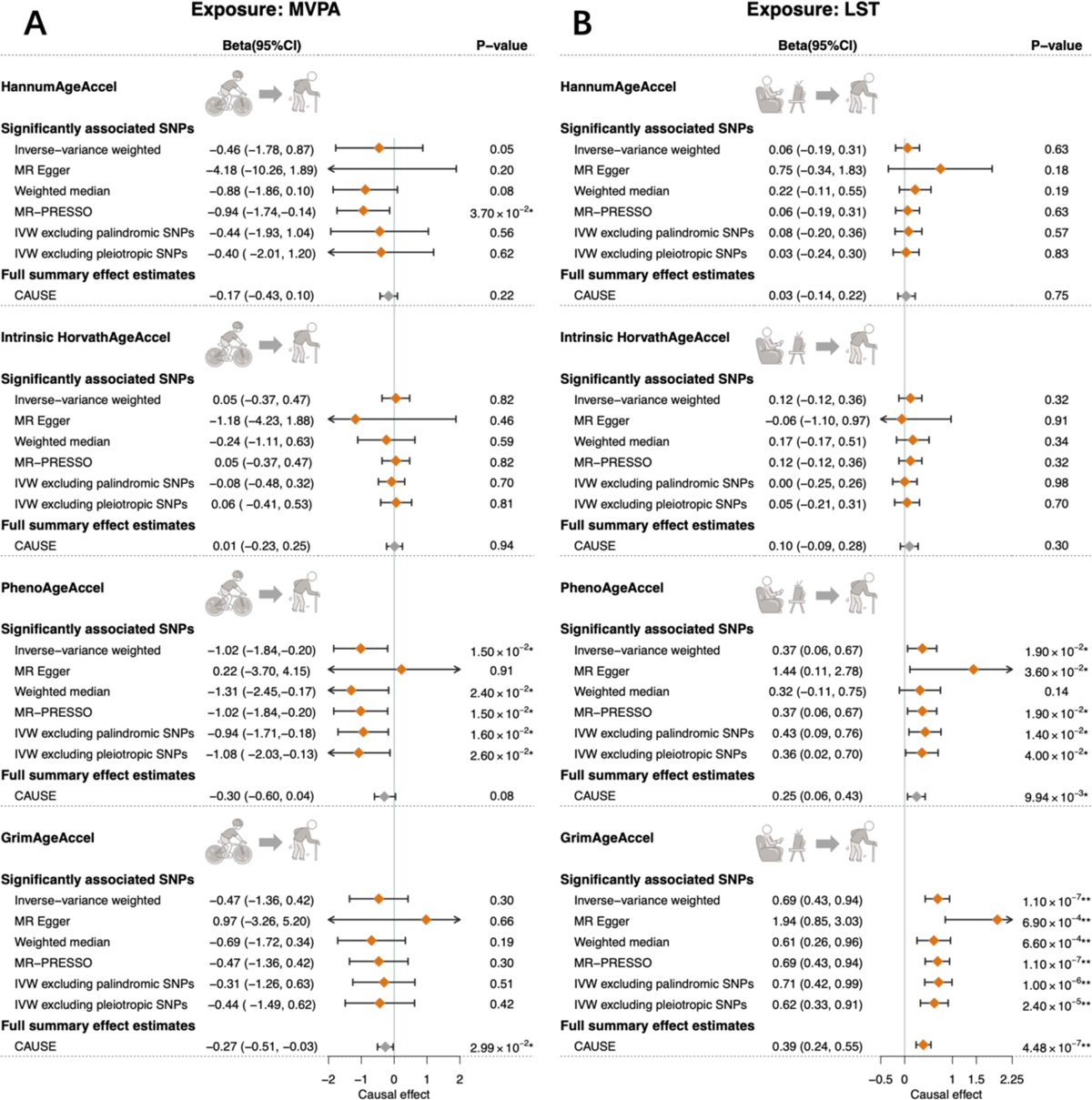
Total effects of physical activity and sedentary behavior on each epigenetic age acceleration using univariable Mendelian randomization. Causal effects of MVPA on EAA are shown in the left panel (A), and causal effects of LST on EAA are shown in the right panel (B). Diamonds represent the point estimates, and error bars represent 95% confidence intervals. Estimates with gray diamond for CAUSE indicate that the null hypothesis for model fitting was not rejected in CAUSE (*P*_causal vs. sharing_ > 0.05), suggesting that the data are more likely to be explained by horizontal pleiotropic effects. One asterisk (*) represents *P* < 0.05 and two asterisks (**) represent the tests that survived Bonferroni correction (*P* < 6.25×10^−3^). IVW (inverse-variance weighted), SNPs (single-nucleotide polymorphisms), MVPA (Moderate-to-vigorous intensity physical activity during leisure time), LST (leisure screen time), HannumAgeAccel (Hannum age acceleration), Intrinsic HorvathAgeAccel (intrinsic Horvath age acceleration), PhenoAgeAccel (PhenoAge acceleration), GrimAgeAccel (GrimAge acceleration), and 95%CI (95% confidence interval).

As for the reverse direction, 10, 26, 11, and 4 SNPs were utilized as IVs for HannumAgeAccel, intrinsic HorvathAgeAccel, PhenoAgeAccel, and GrimAgeAccel, respectively, with *F*-statistics indicating minimal weak instrument bias (all *F*-statistics > 10; **Supplementary Table 3**). No evidence was found to support a reverse effect of any genetically predicted EAA measure on MVPA or LST **(Supplementary Table 12)**.

Leveraging genome-wide summary statistics, evidence from CAUSE further supported the significant causal relationships between LST and GrimAgeAccel (median causal effect = 0.39, 95%CIs = 0.24 ∼ 0.55, *P*_causal vs. sharing_ = 7.00×10^−3^; **Figure 2** and **Supplementary Table 13**). The low absolute value of median shared effect (−0.04) and the low *q* value (0.04) implied that horizontal pleiotropy was limited. CAUSE yielded a suggestively significant estimate for genetically predicted LST with PhenoAgeAccel (median causal effect = 0.25, 95%CIs = 0.06 ∼ 0.43), but not for genetically predicted MVPA with PhenoAgeAccel (median causal effect = −0.30, 95%CIs = −0.60 ∼ 0.04). Nevertheless, results of modeling tests suggested that both associations (LST/MVPA-PhenoAgeAccel) were more likely to be explained by horizontal correlated pleiotropy rather than causality (*P*_causal vs. sharing_ > 0.05).

### Multivariable Mendelian randomization

We prioritized evaluating the independent causal relationship between genetically predicted LST and GrimAgeAccel, based on its robust overall effect confirmed in univariable MR by both the conventional IV-based method and the genome-wide summary statistics-based approach. Taking into account the effect of MVPA, results of Model 1 demonstrated a statistically significant independent effect of genetically predicted LST on GrimAgeAccel (**Figure 3 and Supplementary Table 14)**. Model 2, which incorporated LST with each potential confounder (other than MVPA), provided additional support for its independent causal role. Causal estimates obtained from each separate analysis consistently demonstrated the same direction and were largely statistically significant (except for educational attainment, which yielded to an estimate of suggestive significance; **Figure 3 and Supplementary Table 14**). Model 3 including all potential confounders again generated directionally consistent “LST-GrimAgeAccel” causal associations, despite the attenuated magnitude of effect and the weaker significance. Most conditional *F*-statistics for LST indicated a minimal likelihood of weak instrument bias in multivariable MR settings (>10), with a few exceptions for example when jointly considering MVPA, educational attainment, and BMI **(Supplementary Table 15)**.

**Figure 3.**
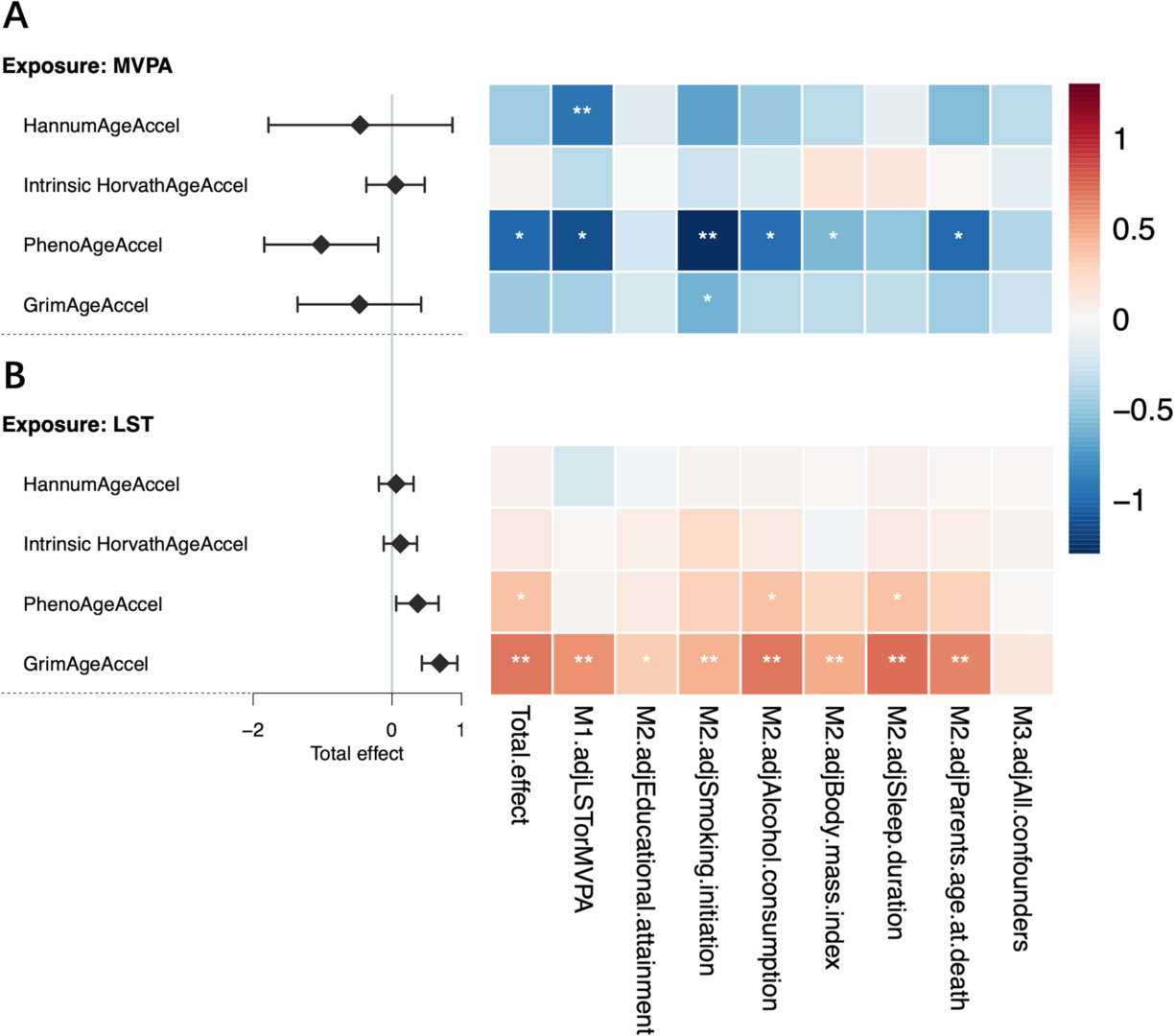
Independent effects of physical activity and sedentary behavior on each epigenetic age acceleration using multivariable Mendelian randomization analysis. Independent causal effects of MVPA on EAA are shown in the upper panel (A), and independent causal effects of LST on EAA are shown in the lower panel (B). One asterisk (*) represents *P* < 0.05 and two asterisks (**) represent the tests that survived Bonferroni correction (*P* < 6.25×10^−3^). Diamonds represent the point estimates of total causal effects, and error bars represent 95% confidence intervals. M1 (Model 1) estimates the independent effects of MVPA and LST from each other. M2 (Model 2) estimates the independent effects of exposures after adjusting for one confounder at a time. M3 (Model 3) estimates the independent effects of exposures after adjusting for all confounders together. Blue represents a negative effect and red represents a positive effect. The darker the color, the larger the absolute value of the effect. MVPA (Moderate-to-vigorous intensity physical activity during leisure time), LST (leisure screen time), HannumAgeAccel (Hannum age acceleration), Intrinsic HorvathAgeAccel (intrinsic Horvath age acceleration), PhenoAgeAccel (PhenoAge acceleration), and GrimAgeAccel (GrimAge acceleration), M2.adjEducational.attainment (causal effects of exposures on outcomes after controlling by educational attainment), M2.adjSmoking.initiation (causal effects of exposures on outcomes after controlling by smoking initiation), M2.adjAlcohol.consumption (causal effects of exposures on outcomes after controlling by alcohol consumption), M2.adjSleep.duration (causal effects of exposures on outcomes after controlling by sleep duration), and M2.adjParents.age.at.death (causal effects of exposures on outcomes after controlling by longevity).

For the “LST-PhenoAgeAccel” and the “MVPA-PhenoAgeAccel” associations that exhibited suggestive significance in univariable MR, the directions of the estimates derived from Models 1-3 remained consistently aligned with their corresponding overall effects. Nevertheless, half of the causal estimates failed to reach suggestive significance, and only the effect of genetically predicted MVPA on PhenoAgeAccel after adjusting for smoking survived Bonferroni corrections (**Figure 3 and Supplementary Table 14**).

### Tissue-partitioned Mendelian randomization

A tissue-partitioned MR was finally conducted to explore whether the “brain-tissue instrumented LST” or the “skeletal muscle-tissue instrumented LST” predominantly drives the observed causal effect of LST on GrimAgeAccel. Among the 130 original IVs for LST, 28 were identified with strong evidence of colocalization for brain-tissue-derived gene expressions, and 30 for skeletal muscle-tissue-derived gene expressions **(Supplementary Table 16)**. The average effect sizes of the two IV sets on LST were virtually identical (brain = 0.028 vs. skeletal muscle = 0.027).

As shown in **Figure 4 and Supplementary Table 17**, univariable MR suggested significant genetically predicted effects of both brain-(*β*_IVW_ = 0.55, 95%CIs = 0.00 ∼ 1.10, *P* = 4.90×10^−2^) and skeletal muscle-tissue (*β*_IVW_ = 0.86, 95%CIs = 0.35 ∼ 1.37, *P* = 9.70×10^−4^) instrumented LST on GrimAgeAccel. Multivariable MR concomitantly incorporating both subcomponents demonstrated a significant effect of skeletal muscle-tissue instrumented LST on GrimAgeAccel (*β*_IVW_ = 1.16, 95%CIs = 0.22 ∼ 2.09, *P* = 1.05×10^−2^). We also repeated the analysis with an expanded set of IVs using a more relaxed clumping strategy of *P* < 1×10^-5^ and an LD window of ± 1.0 Mb (*r^g^*< 0.001), and generated largely consistent results **(Supplementary Table 17-19)**. Unfortunately, the conditional *F*-statistics for tissue-specific instrumented LST were all below 10 (**Supplementary Table 18**).

**Figure 4.**
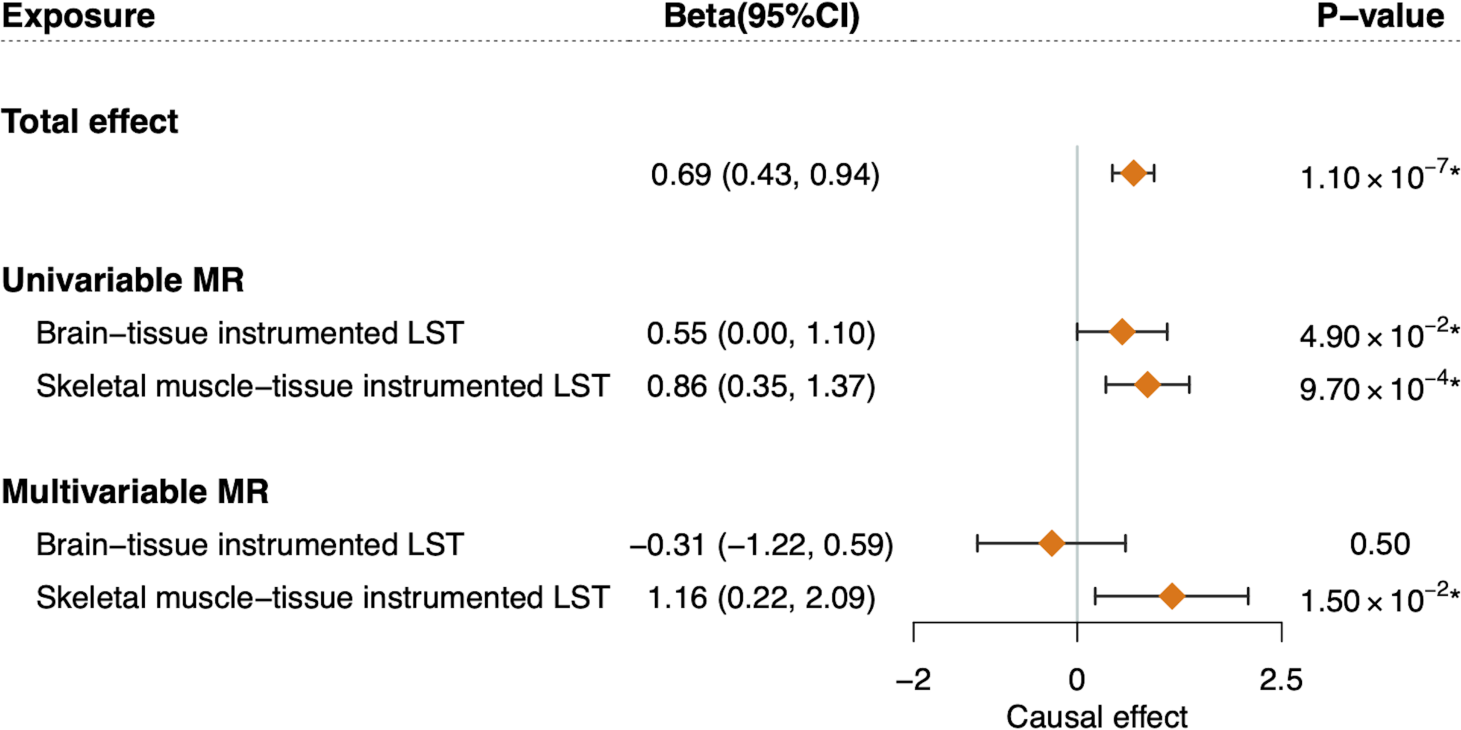
The effects of brain- and skeletal muscle-tissue instrumented LST on GrimAgeAccel using tissue-partitioned Mendelian randomization analysis. Univariable MR analysis evaluates the unadjusted effects of phenotypic subcomponents of brain-tissue instrumented LST and skeletal muscle-tissue instrumented LST on GrimAgeAccel, while multivariable MR estimates the putatively independent effects of each subcomponent with IVs weighted by their PPH4 values for each tissue type. Diamonds represent the point estimates, and error bars represent 95% confidence intervals. One asterisk (*) represents *P* < 0.05. LST (leisure screen time), GrimAgeAccel (GrimAge acceleration), MR (Mendelian randomization), and 95%CI (95% confidence interval).

## Discussion

The current study investigated the shared genetic background and causal relationships between MVPA and LST with four EAA measures, utilizing the largest combination of available datasets and a comprehensive analytical framework. Our findings reveal moderate genetic overlaps of MVPA or LST with both PhenoAgeAccel and GrimAgeAccel. Among these, we highlight strong evidence for an independent causal link between increased LST and accelerated biological aging measured by GrimAgeAccel, with skeletal muscle-related biological pathways playing a predominant role in mediating this effect.

Accumulative studies have consistently shown that the rate of epigenetic aging can be influenced by environmental or lifestyle factors^52^. However, the strength of such relationships varies across different epigenetic clocks, possibly due to the slightly diverse aspects of aging that each clock captures^5^ ^53^. The second-generation clocks, trained on aging-related outcomes rather than solely chronological age, have been found to exhibit stronger correlations with health-related behaviors compared to the first-generation clocks^53^. Multiple observational studies have reported links between higher levels of physical activity with slower PhenoAgeAccel and GrimAgeAccel but not with HannumAgeAccel or intrinsic HorvathAgeAccel^14^ ^15^. Our identification of genome-wide genetic correlations for MVPA or LST with only the acceleration of the second-generation epigenetic clocks provides additional support for prior observational findings from a genetic perspective.

Among the observed genetic overlaps, our MR study further uncovered compelling evidence to support a robust causal effect of genetically predicted LST on GrimAgeAccel, which remains significant even after accounting for the effects of MVPA and other major confounders. In addition, we discerned a marginal association between genetically predicted longer LST with faster PhenoAgeAccel. Two previous cross-sectional studies reported a positive correlation between sedentary time and GrimAgeAccel (*β* = 0.043, *P* = 0.015; *β* = 0.200, *P* = 0.025), with no correlations found for the other EAA measures^14^ ^54^. A more recent study using prospective data revealed an acceleration of GrimAge in the sedentary group compared with the active group (*P* = 0.034), but this association was largely attenuated after adjusting for other lifestyle factors^55^. While these studies hint at a potential link between sedentary behavior and GAA, the evidence is largely weakened by the inherent limitations of the observational design, primarily due to confounding factors and reverse causality, as well as by the single-point measurements of EAA, limiting the tracking of longitudinal changes of epigenetic clock. Utilizing both exposure-associated SNPs and genome-wide summary results, we have for the first time confirmed the causal relationships between LST with GrimAgeAccel, alongside a potential association with PhenoAgeAccel, under a comprehensive MR framework. Our work closely aligns with and greatly expands upon the previous observational studies by providing more reliable evidence of causal inference, thereby paving the way for targeted interventions.

Our MR study also revealed a marginal causal effect of genetically predicted MVPA on PhenoAgeAccel, aligning with a cross-sectional observational study (*β =* −0.26, *P* = 0.021)^14^. In contrast to LST, there has already been one previous MR conducted to examine the causal links between MVPA and two measures of EAA – PhenoAgeAccel (*β*_IVW_ = 0.368, *P* = 0.747) and GrimAgeAccel (*β*_IVW_ = −0.186, *P* = 0.837), which found no significant results^23^. However, the limited number of IVs adopted by this study (6 compared to ours 15), derived from a small-scale MVPA GWAS, might have compromised the precision of estimates due to insufficient power. By using the largest and most updated GWAS summary data for MVPA, findings from our MR suggested an inverse effect of MVPA on PhenoAgeAccel. This effect, however, largely dissipated after accounting for the effects of LST and other confounding factors, calling for further validation with more powerful IVs as they become available. Compared with the robust effects observed for LST, the substantially attenuated associations between MVPA and EAA measures further highlight the primary causal contribution of LST, rather than MVPA, in accelerating epigenetic aging.

Disintegrating genetically predicted LST, the major aging-accelerating risk factor, into tissue-partitioned subcomponents, our work further highlights an important role of skeletal muscle in driving its causal effect on GrimAgeAccel. Loss of muscle mass and strength is a well-established distinctive feature of the aging process^56^, closely related to mobility impairments, physical frailty, and all-cause mortality^57–59^. Several mechanisms have been proposed to explain the alterations in skeletal muscle during aging, such as an imbalance between protein synthesis and degradation^60^, mitochondrial dysfunction^61^, a decrease in type II fiber satellite cells^62^, and infiltration of intramuscular and intermuscular fat^63^. Some of these mechanisms are indeed supported by the detected genes whose skeletal muscle-specific expression was colocalized with LST GWAS associations in our analysis **(Supplementary Table 16)** – for instance, *IGFBP2* (involved in IGF signaling pathways which closely influence protein synthesis and degradation^64^) and *ATP5J2* (involved in mitochondrial function and energy metabolism^65^), indicating energy use and metabolism alterations in muscles during prolonged sedentary periods. A plausible explanation for our identification is that the altered expression of genes associated with LST in skeletal muscle exacerbates muscle atrophy and weakness through multiple pathways (such as a decrease of energy expenditure and metabolism alterations)^66^ ^67^, thereby accelerating biological aging^58^ ^64^. Future research is needed to uncover the exact biological mechanisms, especially those related to muscle wasting or weakness during the aging process.

Taken together, our research provides valuable implications for clinical practice and public health policy by emphasizing the importance of reducing sedentary time as an effective lifestyle intervention to promote healthy aging. Our findings suggest that the acceleration of the biological aging process observed among individuals with a physically inactive lifestyle, resulting from prolonged sedentary time, cannot be fully offset by short periods of exercise or changes in other health-related lifestyles. Therefore, we advocate minimizing the overall sedentary time and adopting “interval activity” and “exercise snacks” strategies, encouraging short breaks for standing, walking, or stretching during prolonged sitting to stimulate muscle activity and mitigate the adverse effects of sedentarism^68^ ^69^. Furthermore, our results emphasize the potential of the second-generation epigenetic clocks, particularly GrimAge, as a valuable tool for assessing the effects of interventions targeting the reduction of sedentary behavior.

Several limitations should be acknowledged. First, using self-reported MVPA as a proxy for physical activity may constrain our results by omitting factors like activity type, duration, intensity, and objective measurements. Additionally, our focus on self-reported LST may not capture the effects of other sedentary behaviors or differentiate between “mentally passive” (such as watching TV) and “mentally active” (such as using computers) sedentary activities due to data limitation^70^. Future investigations should delve deeper into these aspects for a more comprehensive understanding. Second, sample overlap in two-sample MR design is an important issue that needs to be considered^71^. In this study, ∼5% of participants in the exposure

GWASs and ∼20% of participants in the outcome GWASs included several of the same studies (inCHIANTI, TwinsUK, and the Rotterdam Study, etc.). To address this, we used the CAUSE method to correct for the sample overlap, reconfirming the robust causal effect of LST on GrimAgeAccel. Third, although our univariable MR yielded highly robust results, we observed that certain conditional *F*-statistics fell below 10 in multivariable MR settings. This suggests the potential presence of weak instrument bias, which could reduce the power of independent causal estimates in multivariable MR^47^. Therefore, the results of multivariable MR should be interpreted with caution, and future studies should aim to augment sample sizes and IVs to validate our findings. Finally, it should be noted that our tissue-partitioned MR analysis was hypothesis-driven rather than data-driven, indicating that our findings do not exclude the potential roles of other tissue-related pathways (beyond brain or skeletal muscle tissues) in driving the causal effect of genetically predicted LST on GrimAgeAccel. Future research is encouraged to explore these additional mechanisms.

## Conclusion

To conclude, our study provides evidence in support of physically inactive lifestyles, especially increased sedentary time, as a modifiable causal risk factor of epigenetic aging acceleration. Findings from tissue-partitioned analysis shed new light on the underlying mechanisms, with implications for skeletal muscle tissue-related pathways. Our work emphasizes the importance of reducing sedentary time as a preventive strategy to delay the aging process and promote healthy aging.

## Availability of data and codes

The data used in this study are publicly available. GWAS summary data are accessible in all original studies. GWAS summary data of leisure-time physical activity and sedentary behavior, https://www.ebi.ac.uk/gwas/publications/36071172, GWAS summary data of four epigenetic age acceleration measures, https://www.ebi.ac.uk/gwas/publications/34187551 or https://datashare.ed.ac.uk/handle/10283/3645, GWAS summary data of educational attainment, http://www.thessgac.org/data, GWAS summary data of smoking and drinking, https://www.ebi.ac.uk/gwas/publications/30643251, GWAS summary data of BMI, https://zenodo.org/record/1251813, GWAS summary data of sleep duration, https://www.ebi.ac.uk/gwas/publications/30846698, GWAS summary data of parents age at death, https://www.ebi.ac.uk/gwas/publications/29227965.

The data of eQTL are accessible at the website of SMR (Summary Mendelian Randomization, https://cnsgenomics.com/software/smr/), which has already mapped the eQTL data to the hg19 genome build using the GRCh37 reference assembly.

This paper does not report any original code. The software, R packages, and other resources used in this study are accessible at: LDSC, https://github.com/bulik/ldsc, PLINK, https://www.cog-genomics.org/plink/1.9/, LDlinkR, https://github.com/CBIIT/LDlinkR, TwoSampleMR, https://mrcieu.github.io/TwoSampleMR/, MR-PRESSO, https://github.com/rondolab/MR-PRESSO, NHGRI-EBI GWAS Catalog, https://www.ebi.ac.uk/gwas/, CAUSE, https://github.com/jean997/cause, MVMR, https://github.com/WSpiller/MVMR, MendelianRandomization, https://github.com/cran/MendelianRandomization, Coloc, https://chr1swallace.github.io/coloc/.

Any additional information required to reanalyze the data reported in this paper is available from the corresponding author (Xia Jiang, PhD) upon reasonable request.

## Consent for publication

Not applicable.

## Ethics approval and consent to participate

In our study, we utilized publicly available summary-level data from original studies that had received ethical approval and consent for participation.

## Patient and public involvement

Patients and/or the public were not involved in the design, or conduct, or reporting, or dissemination plans of our research.

## Acknowledgments

We thank all the patients, staff, and investigators who contributed to the summary statistics used in this study. The GWAS meta-analysis of physical activity and sedentary behavior was conducted by Wang et al. The GWAS meta-analysis of EAA measures was conducted by McCartney et al. The GWAS summary data of educational attainment from the Social Science Genetic Association Consortium was conducted by Lee et al. The GWAS summary data of smoking and drinking from the GWAS & Sequencing Consortium of Alcohol and Nicotine was conducted by Liu et al. The GWAS meta-analysis of BMI from the UK Biobank and the Genetic Investigation of Anthropometric Traits Consortium was conducted by Pulit et al. The GWAS summary data of sleep duration from the UK Biobank was conducted by Dashti et al. The GWAS summary data of parental longevity from the UK Biobank was conducted by Pilling et al. The meta-analysis of brain eQTL data was conducted by Qi et al., and the skeletal muscle eQTL data was obtained from the GTEx Consortium.

## Equity, diversity, and inclusion statement

We support inclusive, diverse, and equitable conduct of research.

## Author’s contributions

Conceptualization: X.J., J.L., X.Z., X.W.

Data curation: X.Z., X.W.

Formal analysis: X.Z., X.W., L.H., L.S., Y.Q., C.X., C.Q., J.H., Q.D., J.Z.

Funding acquisition: X.J.

Investigation: X.Z., X.W., L.H., R.X.

Methodology: L.H., J.X., R.X., Y.H., M.T.

Project administration: X.Z.,

X.W., S.Z., J.Z., T.Y., B.Y., X.S., T.H.

Supervision: X.J., J.L., M.F., T.Z., J.L.

Writing-original draft: X.Z., X.W.

Writing-review & editing: X.J., X.Z., X.W., L.H.

