## Supplementary File 2 for "Leisure-time physical activity, sedentary behavior, and biological aging: evidence from genetic correlation and Mendelian randomization analyses"

**Supplementary File 2. Details of tissue-partitioned Mendelian randomization analysis**

**Tissue-specific gene expression data**

We obtained the eQTL (expression quantitative trait loci) data for brain from a meta-analysis study (N = 1,194, all of European ancestry) of 10 brain regions (anterior cingulate cortex, caudate basal ganglia, cerebellar hemisphere, cerebellum, cortex, frontal cortex BA9, hippocampus, hypothalamus, nucleus accumbens basal ganglia, and putamen basal ganglia)[1]. We obtained the eQTL data of skeletal muscle from GTEx consortium v8 (N = 706, ~80% European ancestry)[2]. The datasets were both downloaded from the SMR (summary Mendelian randomization) website (https://cnsgenomics.com/software/smr/), which has already mapped the eQTL data to the hg19 genome build using the GRCh37 reference assembly[3].

**Genetic colocalization**

We conducted genetic colocalization using the Bayesian method "*coloc*" to assess whether the same variant is causal in both a GWAS and an eQTL study[4 5]. Colocalization analyses were performed at each locus, encompassing all SNPs located within a 200 kb window around each independent MVPA/LST-associated SNP ($P < 5\times10^{-8}$ and an LD window of ± 1.0 Mb ($r^2$ < 0.001)). To verify the robustness of our results, we also screened an expanded set of IVs using a more relaxed clumping strategy ($P < 1\times10^{-5}$ and an LD window of ± 1.0 Mb ($r^2$ < 0.001)) to repeat our analysis. The colocalization analysis was first conducted with eQTL data derived from brain tissue, and then followed by a separate round with eQTL data from skeletal muscle tissue. The "*coloc*" method provided the posterior probability of five competing hypotheses (PPH0-PPH4). In our study, a locus was considered colocalized if PPH4 ≥ 0.8, indicating both GWAS and eQTL data are associated with a common causal variant[4]. Consequently, the initial set of IVs was divided into two sets of tissue-specific IVs, with each set specifically indexing the "brain-tissue instrumented MVPA/LST" and the "skeletal muscle-tissue instrumented MVPA/LST". To mitigate the potential impact of strong regional linkage disequilibrium (LD) structure, variants within the Major Histocompatibility Complex (MHC) region (chr6: 25M-35M) were excluded in our analysis.

**Mendelian randomization analysis**

Exposure-outcome pairs that showed robust causal relationships in our univariable Mendelian randomization (MR) and directionally consistent estimates in multivariable MR were selected for inclusion in the tissue-partitioned MR analysis. Tissue-partitioned MR was performed in both univariable and multivariable settings. In univariable setting, we first evaluate the unadjusted effects of phenotypic subcomponents of exposures on outcomes, using each set of tissue-specific instrument for the exposures. Then, in multivariable setting, we estimate the putatively genetically predicted effects of exposures instrumented by brain and skeletal muscle tissues independently on outcomes. IVs for the phenotypic subcomponents were weighted by their PPH4 values for each tissue type in multivariable settings, and the model was developed to simultaneously incorporate both sets of tissue-specific instruments. Given the reduced number of IVs and the resulting decrease in statistical power, we used a conventional significance threshold of $P < 0.05$ in the analysis.

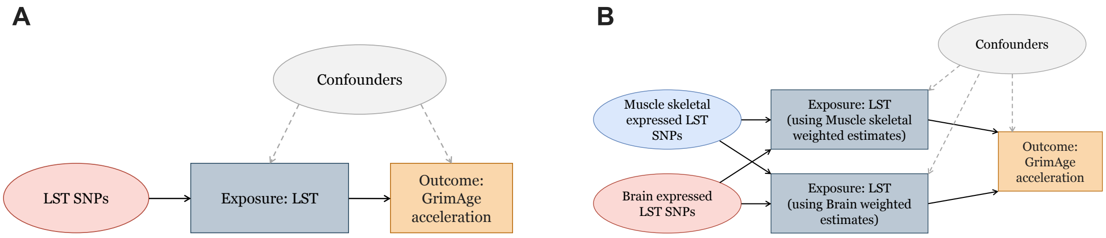


**Supplementary Figure 1. An overview of the tissue-partitioned Mendelian randomization analysis**

Left panel (A) shows the design of a traditional MR analysis, while right panel (B) illustrates the design of a tissue-partitioned MR analysis. In the multivariable setting, the independent effects of phenotypic subcomponents of exposures are estimated by accounting for the effects of other tissues. IVs for the phenotypic subcomponents were weighted by their PPH4 values for each tissue type. LST (leisure screen time), SNP (single nucleotide polymorphisms).
